# COVID infection severity in children under 5 years old before and after Omicron emergence in the US

**DOI:** 10.1101/2022.01.12.22269179

**Authors:** Lindsey Wang, Nathan A. Berger, David C. Kaelber, Pamela B. Davis, Nora D. Volkow, Rong Xu

## Abstract

**Importance:** Pediatric SARS-CoV-2 infections and hospitalizations are rising in the US and other countries after the emergence of Omicron variant. However data on disease severity from Omicron compared with Delta in children under 5 in the US is lacking.

**Objectives:** To compare severity of clinic outcomes in children under 5 who contracted COVID infection for the first time before and after the emergence of Omicron in the US.

**Design, Setting, and Participants:** This is a retrospective cohort study of electronic health record (EHR) data of 79,592 children under 5 who contracted SARS-CoV-2 infection for the first time, including 7,201 infected between 12/26/2021-1/6/2022 when the Omicron predominated (Omicron cohort), 63,203 infected between 9/1/2021-11/15/2021 when the Delta predominated (Delta cohort), and another 9,188 infected between 11/16/2021-11/30/2021 when the Delta predominated but immediately before the Omicron variant was detected in the US (Delta-2 cohort).

**Exposures:** First time infection of SARS-CoV-2.

**Main Outcomes and Measures:** After propensity-score matching, severity of COVID infections including emergency department (ED) visits, hospitalizations, intensive care unit (ICU) admissions, and mechanical ventilation use in the 3-day time-window following SARS-CoV-2 infection were compared between Omicron and Delta cohorts, and between Delta-2 and Delta cohorts. Risk ratios, and 95% confidence intervals (CI) were calculated.

**Results:** Among 7,201 infected children in the Omicron cohort (average age, 1.49 ± 1.42 years), 47.4% were female, 2.4% Asian, 26.1% Black, 13.7% Hispanic, and 44.0% White. Before propensity score matching, the Omicron cohort were younger than the Delta cohort (average age 1.49 vs 1.73 years), comprised of more Black children, and had fewer comorbidities. After propensity-score matching for demographics, socio-economic determinants of health, comorbidities and medications, risks for severe clinical outcomes in the Omicron cohort were significantly lower than those in the Delta cohort: ED visits: 18.83% vs. 26.67% (risk ratio or RR: 0.71 [0.66-0.75]); hospitalizations: 1.04% vs. 3.14% (RR: 0.33 [0.26-0.43]); ICU admissions: 0.14% vs. 0.43% (RR: 0.32 [0.16-0.66]); mechanical ventilation: 0.33% vs. 1.15% (RR: 0.29 [0.18-0.46]). Control studies comparing Delta-2 to Delta cohorts show no difference.

**Conclusions and Relevance:** For children under age 5, first time SARS-CoV-2 infections occurring when the Omicron predominated (prevalence >92%) was associated with significantly less severe outcomes than first-time infections in similar children when the Delta variant predominated.

## Introduction

Pediatric SARS-CoV-2 infections and hospitalizations are rising in the US and other countries after the emergence of Omicron variant^1^. This is especially concerning for children under 5 years old since they are not eligible for COVID-19 vaccines and their low rates of previous SARS-CoV-2 infection also limits their pre-existing immunity^2^. Reports from South Africa^3^, Scotland^4^, and England^5^ showed lower rates of hospitalization following Omicron infection compared with the Delta variant infection. However, the data on disease severity from Omicron in children under 5 is lacking. Here we compared severe clinical outcomes including ED visits, hospitalizations, ICU admissions, and mechanical ventilation use in children under age 5 who contracted SARS-CoV-2 infection for the first time during the period when the Omicron predominated in the US and compared them to those in similar children who first infected when the Delta variant predominated through a retrospective study of a large, geographically diverse database of patient electronic health record (EHR) data in the US.

## METHODS

### Study population

This study used the TriNetX Analytics network platform that contains de-identified EHR data of 90 million unique patients from 63 health care organizations in both inpatient and outpatient settings across the US^6^. TriNetX Analytics provides web-based secure access to patient EHR data from hospitals, primary care, and specialty treatment providers, covering diverse geographic locations, age groups, racial and ethnic groups, income levels, and insurance types. Although the data are fully de-identified, end-users can use built-in statistical functions to perform patient-level data analysis, including cohort selection, propensity-score matching, time trend analysis, outcome research, among others. Because this study only queried statistics of de-identified patient records through web-applications and did not involve retrieval, storage, collection, use, or transmittal of individually identifiable data, Institutional Review Board approval and informed consent was not needed or sought.

The study population comprised of three cohorts of children under age 5 with first time SARS-CoV-2 infections: (a) the Omicron cohort (n = 7,201) – contracted first SARS-CoV-2 infection between 12/26/2021–1/6/2022. The CDC’s national genomic surveillance program reports that Omicron accounted over 92% of all circulating virus variants in the US during the two week period of 12/26/2021–1/6/2022^7^; (b) the Delta cohort (n = 63,203) – contracted first SARS-CoV-2 infection between 9/1/2021–11/15/2021 when Delta was the predominant variant (99.0%)^7^; (c) the Delta-2 cohort (n = 9,188) – contracted first SARS-CoV-2 infection between 11/16/2021– 11/30/2021, immediately before the Omicron variant was detected in the US and when Delta was the predominant variant (99.0%)^7^. This second Delta cohort was created to control for later time periods and shorter window of infection.

The status of SARS-CoV-2 infection was based on the ICD-10 diagnosis code of “COVID-19” (U07.1) or lab-test confirmed presence of “SARS coronavirus 2 and related RNA” (9088). The status of adverse clinical outcomes was based on the Current Procedural Terminology (CPT) relevant codes for ED visits (“Emergency Department Visits”, code 1013711), hospitalizations (“hospital inpatient services”, code: 013659), ICU admissions (“Critical Care Services”, code: 1013729), and mechanical ventilation use (“Respiratory ventilation”, codes: 5A1935Z, 5A1945Z, 5A1955Z, 5A09357, 5A09457, 5A09557).

### Statistical analysis

We tested whether severe clinical outcomes in children in the Omicron cohort differed from those in the Delta cohort. The two cohorts were propensity-score matched (1:1 usinga nearest neighbor greedy matching with a caliper of 0.25 times the standard deviation) for demographics (age, gender, race/ethnicity); adverse socioeconomic determinants of health (assessed by ICD-10 codes “Z55-Z65” for “Persons with potential health hazards related to socioeconomic and psychosocial circumstances”) that include employment, housing, education, and economic circumstances; common medical conditions in children including cancer, congenital heart diseases, asthma, influenza and pneumonia, common cold, asthma, type 1 diabetes, type 2 diabetes, anemia and other blood-related disorders, autistic disorders, overweight (BMI ≥ 85th Percentile for Age), underweight (BMI < 5th Percentile for Age) and COVID-19-related medications^8^ including remdesivir, dexamethasone, hydrocortisone, and tocilizumab (assessed by RxNorm codes).

Severity of clinical outcomes in the propensity-score matched Omicron and Delta cohorts was assessed based on the number of ED visits, hospitalizations, ICU admissions and need for mechanical ventilation in the 3-day time-window that followed from the first day of SARS-CoV-2 infection. Overall risk, risk ratios, and 95% confidence interval (CI) were calculated. The same analyses were performed by comparing Delta-2 to Delta cohorts that were propensity-score matched for the same covariates. All statistical tests were conducted on 1/11/2022 within the TriNetX Analytics Platform with significance set at p-value < 0.05 (two-sided).

## Results

### Patient characteristics

The study population comprised of 79,592 children under age 5 who contracted SARS-CoV-2 infection for the first time between 9/1/2021-1/6/2022, including 7,201 in the Omicron cohort, 63,203 in the Delta cohort, and another 9,188 in the Delta-2 cohort. The characteristics of the Omicron and Delta cohort before and after propensity-score matching are shown in **Table 1**. Compared to children in the Delta cohort, children from the Omicron cohort were younger (average age: 1.47 vs 1.73 years), differed in racial and ethnic compositions, had fewer comorbidities and adverse social determinants of health. After propensity-score matching for variates in the table, the differences between the two cohorts were eliminated. Both Omicron and Delta variant disproportionately infected Black children, especially the Omicron variant.

**Table 1.** Characteristics of the Omicron cohort and the Delta cohort before and after propensity matching. Omicron cohort – children under age 5 years who first contracted SARS-CoV-2 infection between 12/26/2021–1/6/2022 when Omicron was predominant and accounted for over 92% of all variants circulating in the US. Delta cohort – children under age 5 years who first contracted SARS-CoV-2 infection between 9/1/2021–11/15/2021 when Delta was predominant and accounted for 99% of all variants circulating in the US. Race and ethnicity as recorded in the TriNetX EHR database were included because they have been associated with both infection risk and severe outcomes of SARS-CoV-2 infections. P-value – significance between the two cohorts based on two-tailed two-proportion z-test conducted within the TriNetX Network.

|  | Before Matching |  |  | After Matching |  |  |
| --- | --- | --- | --- | --- | --- | --- |
|  | Omicron cohort | Delta Cohort | P-value | Omicron cohort | Delta Cohort | P-value |
| **Total number of infected children under 5** | 7,201 | 63,203 |  | 7,198 | 7,198 |  |
| **Age (years, mean±SD)** | 1.49 ± 1.42 | 1.73 ± 1.41 | $<$ .001 | 1.49 ± 1.42 | 1.48 ± 1.42 | 0.76 |
| **Sex (%)** |  |  |  |  |  |  |
| Female | 47.4 | 46.4 | 0.12 | 47.4 | 47.6 | 0.74 |
| Male | 52.6 | 53.6 | 0.12 | 52.6 | 52.3 | 0.74 |
| **Ethnicity (%)** |  |  |  |  |  |  |
| Hispanic/Latinx | 13.7 | 15.5 | < .001 | 13.7 | 13.8 | 0.81 |
| Not Hispanic/Latinx | 33.0 | 53.2 | < .001 | 33.0 | 33.6 | 0.51 |
| Unknown | 53.3 | 31.3 | < .001 | 53.3 | 52.6 | 0.43 |
| **Race (%)** |  |  |  |  |  |  |
| African American/Black | 26.1 | 20.0 | < .001 | 26.1 | 26.8 | 0.34 |
| Asian | 2.4 | 2.5 | 0.59 | 2.4 | 2.5 | 0.91 |
| White | 44.0 | 49.6 | < .001 | 44.0 | 43.4 | 0.49 |
| Unknown | 27.0 | 27.4 | 0.49 | 27.0 | 26.9 | 0.91 |
| **Adverse Social determinants of health (%)** | 1.4 | 2.6 | < .001 | 1.4 | 1.4 | 0.72 |
| **Comorbidities (%)** |  |  |  |  |  |  |
| Cancer | 1.7 | 2.4 | < .001 | 1.7 | 1.5 | 0.29 |
| Congenital heart diseases | 1.8 | 2.9 | < .001 | 1.8 | 1.6 | 0.31 |
| Type 1 diabetes | 0 | 0.05 | 0.06 | 0 | 0 | - |
| Type 2 diabetes | 0 | 0.03 | 0.14 | 0 | 0 | - |
| Asthma | 1.7 | 3.7 | < .001 | 1.7 | 1.5 | 0.32 |
| Blood Disorders including Anemia, Neutropenia, Coagulation Disorders | 2.4 | 4.6 | < .001 | 2.4 | 2.3 | 0.58 |
| BMI ≥ 95<sup>th</sup> Percentile for Age | 0.5 | 1.1 | < .001 | 0.5 | 0.4 | 0.31 |
| BMI between 85<sup>th</sup>-95<sup>th</sup> Percentile for Age | 0.5 | 1.0 | < .001 | 0.5 | 0.5 | 0.91 |
| BMI < 5<sup>th</sup> Percentile for Age | 0.2 | 0.5 | < .001 | 0.2 | 0.2 | 0.86 |
| Influenza and Pneumonia | 3.4 | 5.0 | < .001 | 3.4 | 3.2 | 0.40 |
| Common cold | 2.2 | 3.8 | < .001 | 2.2 | 1.8 | 0.08 |
| Autistic Disorder | 0.3 | 0.7 | < .001 | 0.3 | 0.3 | 0.66 |
| **COVID-19 therapeutics (%)** |  |  |  |  |  |  |
| Dexamethasone | 14.2 | 13.5 | < .001 | 14.2 | 14.2 | 0.98 |
| Hydrocortisone | 6.1 | 8.2 | < .001 | 6.1 | 5.6 | 0.24 |
| Ibuprofen | 25.6 | 26.2 | 0.27 | 25.5 | 25.7 | 0.79 |
| Prednisone | 0.2 | 0.3 | 0.12 | 0.2 | 0.1 | 0.53 |
| Methylprednisolone | 6.8 | 4.3 | < .001 | 6.8 | 6.4 | 0.33 |
| Prednisolone | 5.7 | 6.2 | 0.09 | 5.7 | 5.1 | 0.13 |
| Naproxen | 0.0 | 0.03 | 0.13 | 0.0 | 0.0 | - |
| Tocilizumab | 0.0 | 0.02 | 0.24 | 0.0 | 0.0 | - |

Among 520,624 children under age 5 in the TriNetX database who had any medical encounter with healthcare systems between 9/1/2021-1/6/2022, 14.6% were black, which is significantly lower than the 26.1% among Omicron-infected children (P <0.001) and 20.0% among Delta-infected children (P <0.001), suggesting the disparity in infection with both Delta and Omicron, even in children under 5 years old.

### Lower risks of severe clinical outcomes in the Omicron cohort than in the matched Delta cohort

The risks for severe clinical outcomes in the 3-day time-window following initial SARS-CoV-2 infection in the Omicron cohort (n=7,198) were significantly lower than in the 1:1 matched Delta cohort (n=7,198): ED visits: 18.83% vs. 26.67% (risk ratio or RR: 0.71 [0.66-0.75]); hospitalizations: 1.04% vs. 3.14% (RR: 0.33 [0.26-0.43]); ICU admissions: 0.14% vs. 0.43% (RR: 0.32 [0.16-0.66]); mechanical ventilation: 0.33% vs. 1.15% (RR: 0.29 [0.18-0.46]) (**Figure 1**, top). Control studies comparing the matched Delta-2 and Delta cohorts show no difference (**Figure 1**, bottom), suggesting that the differences in disease severity between Omicron and Delta cohorts are largely attributed to inherent properties of the virus variant itself.

**Figure 1.**
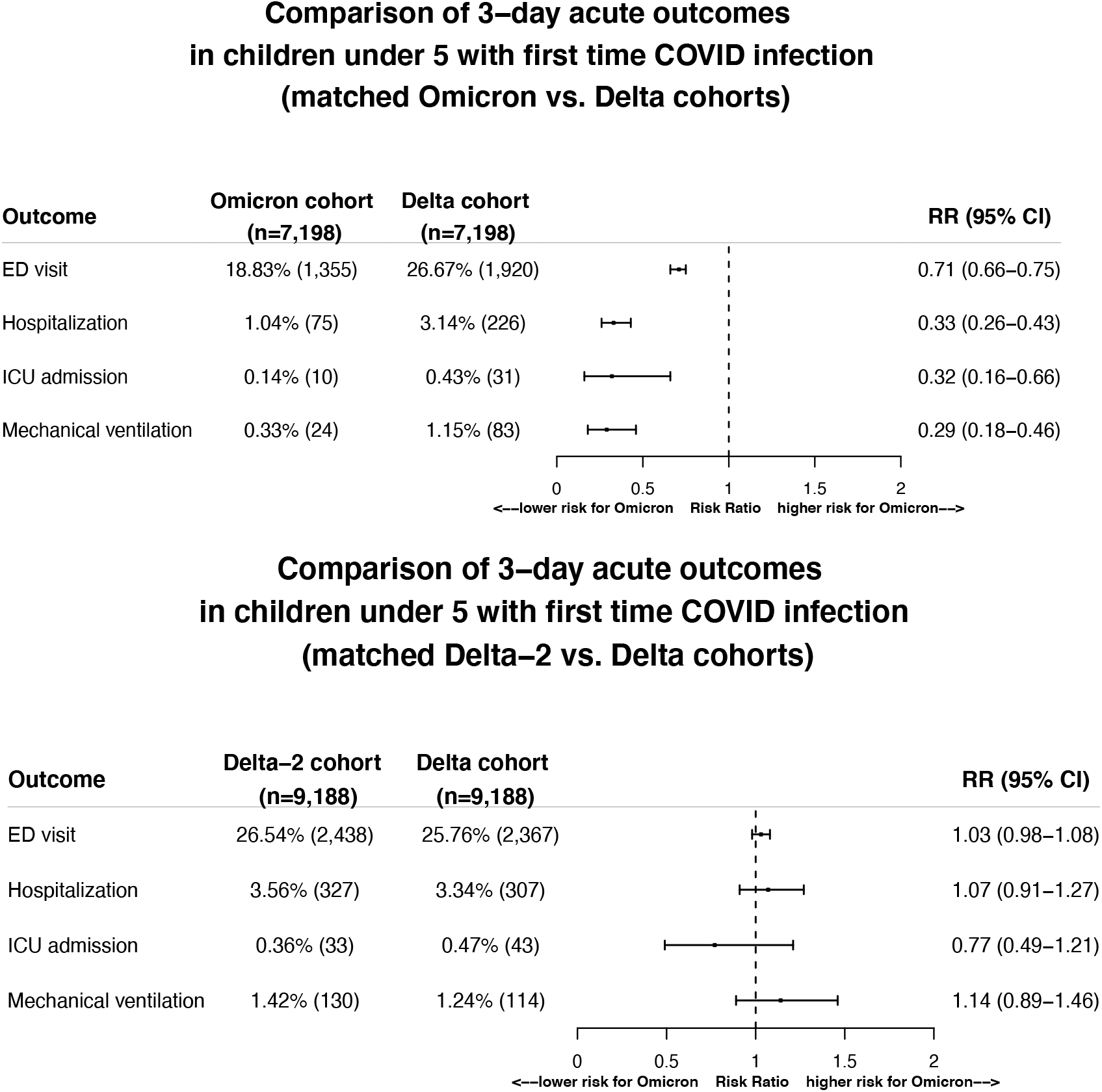
Comparison of severity of clinical outcomes including ED visits, hospitalizations, ICU admissions, mechanical ventilation in the 3-day time-window that followed from the first day of SARS-CoV-2 infection between the matched Omicron and Delta cohorts (Top panel) and between the two matched Delta cohorts (Bottom panel). Omicron cohort – children under 5 who first contracted SARS-CoV-2 infection between 12/26/2021–1/6/2022 when Omicron was predominant and accounted for >92% of all variants circulating in the US. Delta cohort – children under 5 who first contracted SARS-CoV-2 infection between 9/1/2021–11/15/2021 when Delta was predominant and accounted for >99% of all variants circulating in the US. Delta-2 cohort – children under 5 who first contracted SARS-CoV-2 infection between 11/16/2021–11/30/2021, right before the Omicron variant emerged in the US and when Delta accounted for >99% of all variants circulating in the US. Cohorts were propensity-score matched for demographics (age, gender, race/ethnicity), socioeconomic factors, pediatric health conditions, and COVID-19 related medications.

## Discussion

In this study using a nation-wide database of EHRs in the US, children under 5 who were infected with SARS-CoV-2 for the first time when the Omicron variant predominated (>92% of circulating variants) were younger and had fewer adverse health conditions than those infected when the Delta variant predominated. After matching for age, gender, race/ethnicity, adverse socio-economic determinants of health, comorbidities and medications, the risks for ED visits, hospitalizations, ICU admissions and mechanical ventilation within 3 days of infection were significantly lower in children from the Omicron cohort than those from the matched Delta cohort, with 29% reduction in ED visits, 67% reduction in hospitalizations, 68% reduction in ICU admissions, and 71% reduction in mechanical ventilation. Mortality risks were not reported in this study because so few deaths occurred within 3 days of infection in both cohorts. Propensity score matching and the comparison between Omicron and Delta cohorts suggested that the milder disease severity for Omicron were not confounded by factors such as age, race/ethnicity, overall health of those infected, socioeconomic factors, and medications. No differences in outcomes were observed for SARS-CoV-2 infections occurring during the two-week Delta variant period (11/16/2021–11/30/2021) that immediately preceded the emergence of the Omicron variant and the 10-week Delta variant period before it (9/1/2021–11/15/2021), which suggested that findings were not confounded by other factors such as timing, colder weather, and holiday seasons. Since vaccines are not available to children under 5 and the study population comprised of children who had no previous infections by coronavirus, there were no confounding effects of vaccination or prior infections, so there should be no effects of pre-existing immunity. Taken together, these results suggest that the Omicron variant is inherently milder than the Delta variant in infected children under 5.

Our study has several limitations: First, the observational, retrospective nature of this study of patient EHR data could introduce case selection biases, over representation of symptomatic testing, reporting and follow up issues. However, because we compared the different cohort populations all from the TriNetX dataset, these issues should not significantly affect the relative risk analyses. Second, patients in the TriNetX EHR database are those who had medical encounters with healthcare systems contributing to the TriNetX Platform and do not necessarily represent the entire US population. Therefore, results from the TriNetX platform need to be validated in other populations. Again, the observed significant differences in COVID infection severity should hold in other populations as they were all obtained from the TriNetX dataset. Third, both the Omicron and Delta cohorts in our study were defined based on CDC’s national genomic sequence surveillance. Therefore, the Omicron cohort likely contained a few infections with the Delta variant. However, our findings of reduced disease severity in the Omicron cohort compared to the Delta cohort are consistent with findings from Africa^3^, Scotland^4^, and England^5^ that were based on genomic sequences, though this study specifically focused on children under 5 years of age. The fact that there were no differences between the two Delta cohorts yet significant differences between the Omicron and Delta cohorts further suggest that the differences resulted from inherent properties of the variant itself. Finally, the findings apply only to infections that occurred in the US between 12/26/2021– 1/6/2022 when Omicron accounted for more than 92% of all variants. Given the potential for further viral mutations, future studies are imperative to closely follow up disease severity as well as longer term outcomes associated with infections from Omicron or other variants.

In conclusion, our analysis indicates that COVID-19 infections from the Omicron variant in children under 5 years of age were associated with less severe outcomes as compared to the Delta variant period that preceded it. Despite this encouraging result, further studies are needed to monitor the longer-term acute consequences from Omicron infection, the propensity for development of “long COVID”, the rapidity of spread, potential for mutation, and how prior infections alter clinical responses. Additionally, although infections from the Omicron variant, based on this analysis, appear to be milder, because of Omicron’s increased transmissibility, the overall number of ED visits, hospitalizations, ICU admissions, and mechanical ventilator use in children may still be greater with the Omicron variant than the Delta variant.

## Data Availability

All data produced in the present work are contained in the manuscript

## Contributors

RX conceived, designed, and supervised the study, and drafted the manuscript. LW conducted the experiments, performed data analysis and prepared tables and figures. NAB, PBD, DCK, and NDV critically contributed to study design, result interpretation and manuscript preparation. We confirm the originality of content.

## Acknowledgments

We acknowledge support from National Institute on Aging (grants nos. AG057557, AG061388, AG062272), National Institute on Alcohol Abuse and Alcoholism (grant no. R01AA029831), National Institute on Drug Abuse (grant no. UG1DA049435), the Clinical and Translational Science Collaborative (CTSC) of Cleveland (grant no. 1UL1TR002548-01), National Cancer Institute Case Comprehensive Cancer Center (R25CA221718, P30 CA043703, P20 CA2332216).

## Meeting Presentation

No

## References

1. Cloete J, Kruger A, Masha M, et al. Rapid rise in paediatric COVID-19 hospitalisations during the early stages of the Omicron wave, Tshwane District, South Africa. bioRxiv. Published online December 21, 2021. doi:10.1101/2021.12.21.21268108

2. Ledford H. How severe are Omicron infections? Nature. 2021;600(7890):577–578.

3. Wolter N, Jassat W, Walaza S, et al. Early assessment of the clinical severity of the SARS-CoV-2 Omicron variant in South Africa. bioRxiv. Published online December 21, 2021. doi:10.1101/2021.12.21.21268116

4. Sheikh A, Kerr S, Woolhouse M, McMenamin J, Robertson C. Severity of Omicron variant of concern and vaccine effectiveness against symptomatic disease: national cohort with nested test negative design study in Scotland. Published online December 22, 2021. Accessed December 23, 2021. https://www.pure.ed.ac.uk/ws/files/245818096/Severity_of_Omicron_variant_of_concern_and_vaccine_effectiveness_against_symptomatic_disease.pdf

5. Report 50 - Hospitalisation risk for Omicron cases in England. Accessed December 23, 2021. https://www.imperial.ac.uk/mrc-global-infectious-disease-analysis/covid-19/report-50-severity-omicron/

6. TriNetX. Published September 8, 2021. Accessed December 17, 2021. https://trinetx.com/

7. CDC. COVID Data Tracker. Accessed December 23, 2021. https://covid.cdc.gov/covid-data-tracker/#variant-proportions

8. Hospitalized Adults: Therapeutic Management. Accessed December 24, 2021. https://www.covid19treatmentguidelines.nih.gov/management/clinical-management/hospitalized-adults--therapeutic-management/

